# Cefazolin versus Antistaphylococcal Penicillins for *Methicillin-Susceptible Staphylococcus aureus* Bacteremia: A Systematic Review and Meta-Analysis of Randomized Controlled Trials

**DOI:** 10.64898/2026.10.12.26365331

**Authors:** Frederico Larré Pozzebon, Gustavo Treptow Cabral, Giovani Michelin Pacheco, Bibiana Vieira Dias, Rodrigo Serra Macedo, Adriano Martimbianco de Assis

## Abstract

**Purpose:** To compare the efficacy and safety of intravenous cefazolin versus antistaphylococcal penicillins for the treatment of MSSA bacteremia in adults using evidence exclusively from randomized controlled trials.

**Methods:** We searched 5 databases for RCTs comparing definitive intravenous cefazolin with ASPs (oxacillin, cloxacillin, flucloxacillin, nafcillin) in adults with MSSA bacteremia. Primary outcomes were 90-day all-cause mortality and acute kidney injury (AKI). Secondary outcomes included adverse event-related treatment discontinuation and persistent bacteremia (≥72h). Dichotomous outcomes were synthesized as risk ratios (RRs) with 95% confidence intervals (CIs) using random-effects models with Paule-Mandel estimation and Hartung-Knapp-Sidik-Jonkman inference. Fixed-effect Mantel-Haenszel models were performed as sensitivity analyses.

**Results:** Two RCTs randomized 1,656 participants (SNAP, n = 1,341; CloCeBa, n = 315), of whom 1,579 contributed to the 90-day mortality analysis. There was no statistically significant difference in 90-day mortality (RR 0.90, 95% CI 0.57-1.41; I^2^ = 0%). Both trials favoured cefazolin for acute kidney injury, but the random-effects estimate was highly imprecise (RR 0.26, 95% CI 0.00-939,970.59; I^2^ = 81.7%). Treatment discontinuation also favoured cefazolin directionally, although the random-effects estimate was imprecise (RR 0.24, 95% CI 0.00-24.41; I^2^ = 36.2%). Microbiological outcomes were synthesized narratively because definitions and assessment time points differed between trials.

**Conclusion:** Randomized evidence supports cefazolin as a potential definitive treatment option when adequate source control, appropriate antimicrobial dosing, and clinical monitoring can be achieved. However, the limited number of trials and imprecision in pooled estimates warrant cautious interpretation.

## 1. INTRODUCTION

Methicillin-susceptible *Staphylococcus aureus* (MSSA) bacteremia is a serious infection associated with substantial morbidity and 90-day all-cause mortality historically estimated at 15%-20% [1,2]. Intravenous antistaphylococcal penicillins (ASPs), including oxacillin, cloxacillin, flucloxacillin, and nafcillin, have traditionally been standard therapy [3,4]. However, their use is limited by treatment-emergent toxicity, including acute kidney injury (AKI), hepatotoxicity, hypersensitivity, and hematological adverse events, with the dominant toxicity profile varying by agent [5–8]. Cefazolin offers practical advantages, including less frequent dosing and a potentially more favorable safety profile [6,10]. Observational evidence has also associated cefazolin with fewer adverse-event-related treatment discontinuations, although confounding limits causal interpretation [9].

Concerns about the cefazolin inoculum effect (CzIE), an in vitro increase in the minimum inhibitory concentration of cefazolin at high bacterial inocula, have historically limited its use in severe MSSA infections [11–14]. However, the clinical relevance of this phenomenon remains uncertain [13,15,16]. Recent randomized controlled trials (RCTs), including SNAP [3] and CloCeBa [4], provide direct comparative evidence on the efficacy and safety of cefazolin versus ASPs. Nevertheless, differences in outcome definitions, assessment time points, and analytical approaches complicate interpretation across trials.

Although a recent review summarized these results, it was not designed to provide a formal quantitative synthesis of evidence from randomized controlled trials [17]. We therefore conducted a systematic review and meta-analysis restricted to RCTs to compare cefazolin with ASPs for adult MSSA bacteremia. We assessed 90-day all-cause mortality and AKI as the primary efficacy and safety outcomes, respectively, alongside treatment discontinuation and microbiological outcomes. We also examined between-trial heterogeneity, the robustness of findings to analytical assumptions, and the certainty of the evidence using GRADE.

## 2. MATERIALS AND METHODS

### 2.1. Protocol and Registration

This systematic review and meta-analysis followed PRISMA 2020 [18] and the Cochrane Handbook [19]. The protocol was prospectively registered in PROSPERO (CRD420261465101).

### 2.2. Eligibility Criteria and Outcome Definitions

Eligibility criteria were established according to the PICOS framework. We included prospective randomized controlled trials (RCTs), including parallel-group and adaptive trial designs, enrolling adults aged ≥18 years with methicillin-susceptible *Staphylococcus aureus* (MSSA) bacteremia confirmed by at least one positive blood culture. Eligible interventions were intravenous cefazolin administered as definitive therapy or as the randomized initial treatment strategy, compared with intravenous antistaphylococcal penicillins (ASPs), including oxacillin, cloxacillin, flucloxacillin, or nafcillin. Observational studies, non-randomized or quasi-experimental studies, single-arm cohorts, case series, and reviews were excluded.

The primary efficacy outcome was all-cause mortality at 90 days, selected because it was clinically relevant, objectively ascertainable, and reported at a common time point across the included trials. Mortality at 30 days was also specified in the registered review as a secondary time point. We sought data at the prespecified time point and did not substitute mortality measured at a different time point when an exact 30-day estimate was unavailable. The primary safety outcome was acute kidney injury (AKI), using the trial-specific standardized laboratory criteria and assessment periods reported by each study.

Secondary outcomes included treatment discontinuation due to adverse events, persistent bacteremia, late microbiological failure or bacteremia relapse, and length of hospital stay, as specified in the review registration. Persistent bacteremia was defined as continued MSSA-positive blood cultures at least 72 hours after initiation of active therapy, while retaining trial-specific definitions where these differed. Microbiological outcomes were synthesized narratively when operational definitions or assessment time points were not sufficiently comparable for quantitative pooling. For outcomes specified in the registration but not reported in a compatible form by the included trials, no pooled estimate was calculated.

### 2.3. Search Strategy and Study Selection

Searches were conducted in PubMed/MEDLINE, Embase, CENTRAL, Web of Science, and Scopus from inception to July 2026 without restrictions. Clinical trial registries, including ClinicalTrials.gov, were also searched for unpublished or ongoing RCTs. Search strategies combined controlled vocabulary and keywords for cefazolin, MSSA bacteremia, and antistaphylococcal penicillins with validated RCT filters. Complete database-specific strategies, including the search terms and applicable limits, are provided in **Supplementary Table S1**. The search methods were expanded beyond the sources listed in the original PROSPERO registration to include Cochrane CENTRAL and Web of Science and to incorporate clinical trial registry searches. These changes were made to improve retrieval of potentially eligible evidence and are reported transparently as amendments to the registered search plan. Reference lists of eligible trials and relevant reviews were also hand-searched. Records were imported into Rayyan for deduplication and screening. Two reviewers independently screened titles and abstracts and assessed potentially eligible full-text reports. Disagreements were resolved by consensus or adjudication by a third reviewer.

### 2.4. Data Extraction and Risk of Bias Assessment

Two authors independently extracted data using a standardized form. Extracted variables included study characteristics, baseline demographic and clinical features, dosing regimens, outcome definitions and assessment periods, and the exact event counts and denominators reported for efficacy and safety outcomes. Data were extracted according to the analysis populations and outcome-specific denominators reported by each trial; denominators were not assumed to be identical across outcomes.

Methodological quality and risk of bias were assessed independently in duplicate using the Cochrane Risk of Bias 2 (RoB 2) tool [20]. Five core domains were examined: randomisation process, deviations from intended interventions, missing outcome data, measurement of the outcome, and selection of the reported results.

### 2.5. Statistical Analysis and Certainty of Evidence

Dichotomous outcomes were pooled as risk ratios (RRs) with 95% confidence intervals (CIs). Although the registration specified random-effects synthesis (DerSimonian-Laird or restricted maximum likelihood), the final analyses used the Paule-Mandel estimator for between-study variance with Hartung-Knapp-Sidik-Jonkman (HKSJ) inference [21]. The random-effects framework was retained, but the estimator and inferential method differed from those specified in the registration. This decision was made during the analysis and is reported transparently. Fixed-effect Mantel-Haenszel models were additionally conducted as sensitivity analyses to assess the robustness of findings to the choice of meta-analytic model.

Statistical heterogeneity was assessed using Cochran’s Q test and the I^2^ statistic. Given the small number of contributing trials, these measures and the between-study variance were interpreted cautiously and alongside the direction and magnitude of individual trial estimates. Study-specific weights were added to Supplementary Table S2. To explore potential clinical and methodological sources of heterogeneity in AKI outcomes, trial-specific definitions, ascertainment procedures, assessment periods, and management characteristics relevant to AKI were compared descriptively. Sensitivity analyses examined alternative effect measures, including risk differences and odds ratios where appropriate. The robustness of the 90-day mortality findings to missing outcome data was assessed using deterministic best-case, worst-case, and extreme-case assumptions. Analyses were performed in R version 4.4.2 using the *meta* and *metafor* packages [22–24]. Certainty of evidence was assessed using GRADE, considering risk of bias, inconsistency, indirectness, and imprecision [25].

Microbiological endpoints differed operationally: SNAP assessed treatment failure after Day 14, whereas CloCeBa assessed bacteremia relapse through Day 90. Outcomes reported in only one trial or measured differently across trials were not pooled. These outcomes were synthesized narratively to complement the quantitative findings.

## 3. RESULTS

### 3.1. Search Results and Trial Characteristics

The systematic search across five electronic databases identified 572 records: PubMed (n = 22), Web of Science (n = 56), Embase (n = 24), Cochrane CENTRAL (n = 47), and Scopus (n = 423). An additional 11 records were retrieved from ClinicalTrials.gov (n = 11), yielding a total of 583 records. Following automated and manual deduplication, 482 unique records were screened **(Figure 1)**.

**Figure 1.**
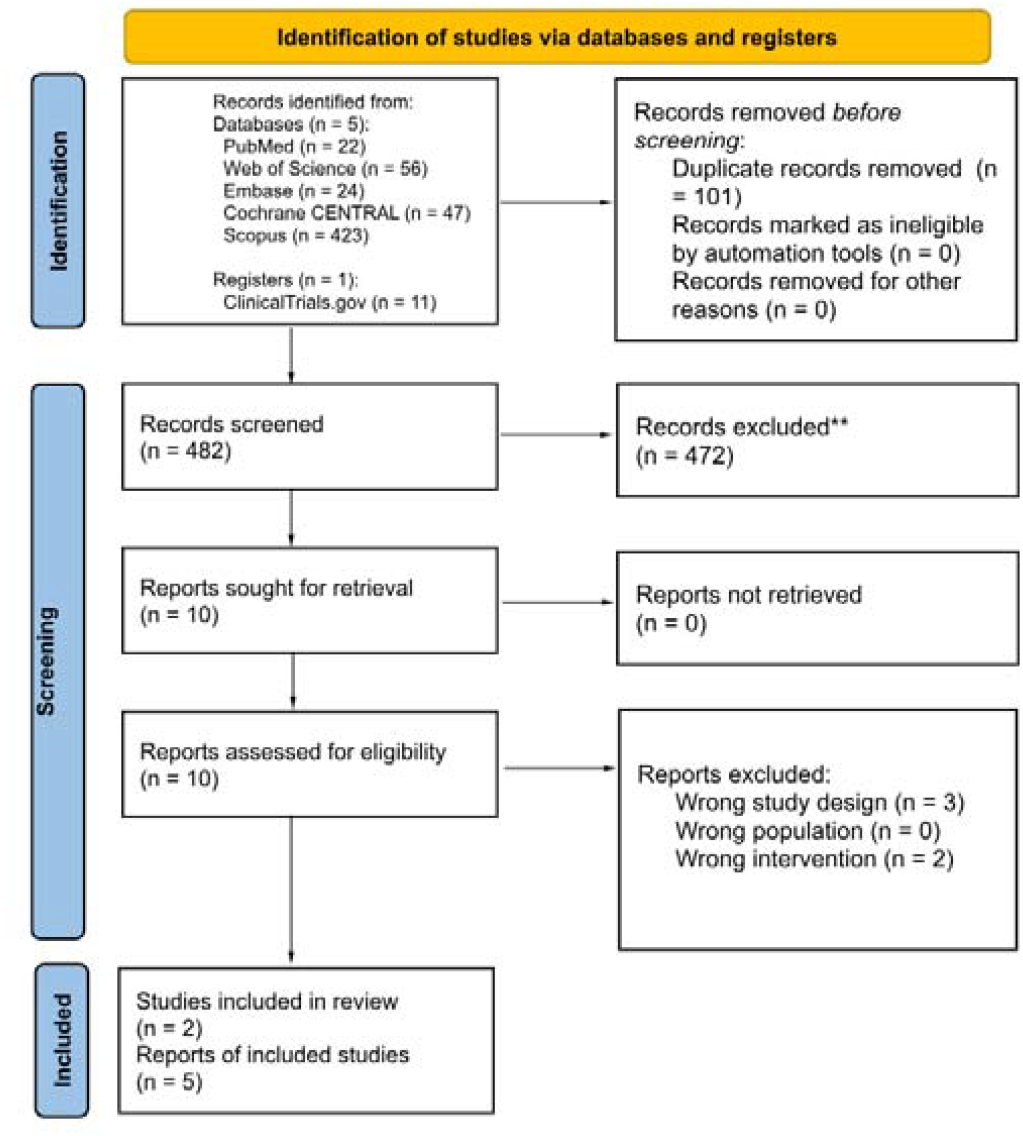
PRISMA flow diagram of the identification, screening and inclusion of randomized controlled trials comparing cefazolin with antistaphylococcal penicillins in adults with methicillin-susceptible *Staphylococcus aureus* bacteremia.

Full-text assessment resulted in two eligible multicenter RCTs: SNAP (n = 1,341 randomized) and CloCeBa (n = 315 randomized) [3,4], for a total randomized population of 1,656 participants. For the 90-day mortality analysis, 1,579 participants contributed data (cefazolin: n = 791; ASPs: n = 788). Outcome-specific denominators varied according to the analysis populations reported by the individual trials. Baseline characteristics were broadly balanced (Table 1).

**Table 1.** Participant characteristics and methodological features of randomized controlled trials comparing cefazolin with antistaphylococcal penicillins in adults with methicillin-susceptible *Staphylococcus aureus* bacteremia.

| Characteristic / Parameter | SNAP Trial (2026) | CloCeBa Trial (2025) |
| --- | --- | --- |
| Primary Reference | Lee et al. (*N Engl J Med*) | Burdet et al. (*Lancet*) |
| Trial Registration | **NCT05137119** | **NCT03248063** |
| Study Design | International, Bayesian adaptive platform, open-label, randomized controlled non-inferiority trial | Prospective, multicenter, open-label, randomized controlled non-inferiority trial |
| Geographic Setting | 91 clinical centers across 8 countries | 21 university and tertiary hospital centers across France |
| Enrollment Period | **February 17, 2022 to August 7, 2024** | **September 5, 2018 to November 16, 2023** |
| Analyzed Population (N) | **1,341 randomized** (671 cefazolin; 670 (flu)cloxacillin); primary mortality analysis: **1,287 evaluable** (645; 642), excluding 54 participants with missing 90-day mortality data; additional protocol-adherent analysis. | **315 randomized** (158 cefazolin; 157 cloxacillin); **ITT/mITT: 292** (146 per group; identical populations); **per-protocol: 223** (115; 108). |
| Age (years), Mean ± SD | **61.0 ± 16.5** | **62.7 ± 16.4** |
| Male Sex, n (%) | **882 / 1,287 (68.5%)** | **215 / 292 (73.6%)** |
| Intervention Arm | Cefazolin: **2 g IV every 8 h**, with renal dose adjustment according to protocol | Cefazolin: **25-50 mg/kg IV every 8 h for the first 7 days**, with subsequent therapy at investigator discretion |
| **Comparator Arm (ASP)** | Cloxacillin 2 g IV every 4 h or flucloxacillin 2 g IV every 4-6 h | Cloxacillin: **25-50 mg/kg IV every 4-6 h for the first 7 days**, with subsequent therapy at investigator discretion |
| **Primary Infection Foci, n (%)** | Vascular catheter/device: ~27%; skin and soft tissue: ~22%; bone and joint: ~18%; infective endocarditis: ~10%; primary/unknown/other: ~23% | Skin and soft tissue: **88/292 (30.1%)**; catheter-related: **64/292 (21.9%)**; bone and joint: **51/292 (17.5%)**; infective endocarditis: **38/292 (13.0%)**; unknown focus: **35/292 (12.0%)** |
| **Key Eligibility Criteria** | Adults ≥18 years with blood culture-confirmed MSSA bacteremia requiring targeted intravenous therapy; key exclusions included severe β-lactam hypersensitivity, imminent death, or more than 72 h of effective therapy before randomisation | Adults ≥18 years with blood culture-confirmed MSSA bacteremia; key exclusions included CNS infection, severe renal impairment, and pregnancy or breastfeeding |
| **Antimicrobial Duration** | ≥14 days total, guided by infection source and clinical protocol | ≥14 days total; treatment after the first 7 days was determined according to clinical circumstances and investigator discretion |
| **Primary Study Endpoint** | **All-cause mortality at 90 days after enrollment**; Bayesian non-inferiority assessment based on the posterior probability | **90-day composite therapeutic success**, incorporating survival, microbiological clearance, absence of microbiological relapse, and clinical success; non-inferiority margin of **12 percentage points** |

**Abbreviations:** ASP, antistaphylococcal penicillins; CNS, central nervous system; ITT, intention-to-treat; IV, intravenous; mITT, modified intention-to-treat; MSSA, methicillin-susceptible *Staphylococcus aureus*; RCT, randomized controlled trial; SD, standard deviation.

### 3.2. Risk of Bias Assessment (Cochrane RoB 2)

For 90-day all-cause mortality, SNAP was judged at low overall risk of bias, whereas the CloCeBa trial presented some concerns (detailed in Table 2 and Supplementary Table S4). Randomisation and allocation concealment were appropriate in both trials. Because 90-day mortality is a hard, objective clinical endpoint, it was considered unlikely to be substantially influenced by the open-label design. For secondary safety outcomes, both trials received some concerns or high risk ratings, mainly because open-label treatment decisions could influence the ascertainment of adverse events, hydration protocols, or decisions to alter/discontinue therapy.

**Table 2.** Domain-specific and overall risk-of-bias assessments for 90-day all-cause mortality and safety outcomes in randomized controlled trials comparing cefazolin with antistaphylococcal penicillins for methicillin-susceptible *Staphylococcus aureus* bacteremia, using the Cochrane RoB 2 tool.

| Study and outcome | D1 | D2 | D3 | D4 | D5 | Overall |
| --- | --- | --- | --- | --- | --- | --- |
| SNAP (2026) - Mortality | Low | Low | Low | Low | Low | **Low** |
| SNAP (2026) - AKI | Low | Low | Low | Some concerns | Low | **Some concerns** |
| SNAP (2026) - Treatment discontinuation due to AE | Low | Low | Low | **High** | Low | **High** |
| CloCeBa (2025) - Mortality | Low | **Some concerns** | **Some concerns** | Low | Low | **Some concerns** |
| CloCeBa (2025) - AKI | Low | **Some concerns** | **Some concerns** | **Low** | Some concerns | **Some concerns** |
| CloCeBa (2025) - Treatment discontinuation due to AE | Low | Some concerns | Low | **High** | Low | **High** |

**Abbreviations:** AKI, acute kidney injury; RoB 2, Cochrane risk-of-bias tool for randomized trials; D1, bias arising from the randomisation process; D2, bias due to deviations from intended interventions; D3, bias due to missing outcome data; D4, bias in measurement of the outcome; D5, bias in selection of the reported result.

### 3.3. Quantitative Synthesis of Clinical Outcomes

#### 3.3.1. Primary Efficacy Outcome: 90-Day All-Cause Mortality

All-cause mortality at 90 days was similar between treatment arms, with no evidence of a survival advantage for either cefazolin or antistaphylococcal penicillins and no statistical heterogeneity across trials **(Figure 2A, Table 3)**. Sensitivity analyses using best-case, worst-case, and extreme-case assumptions for missing outcome data yielded consistent findings **(Table 4)**.

**Table 3.**
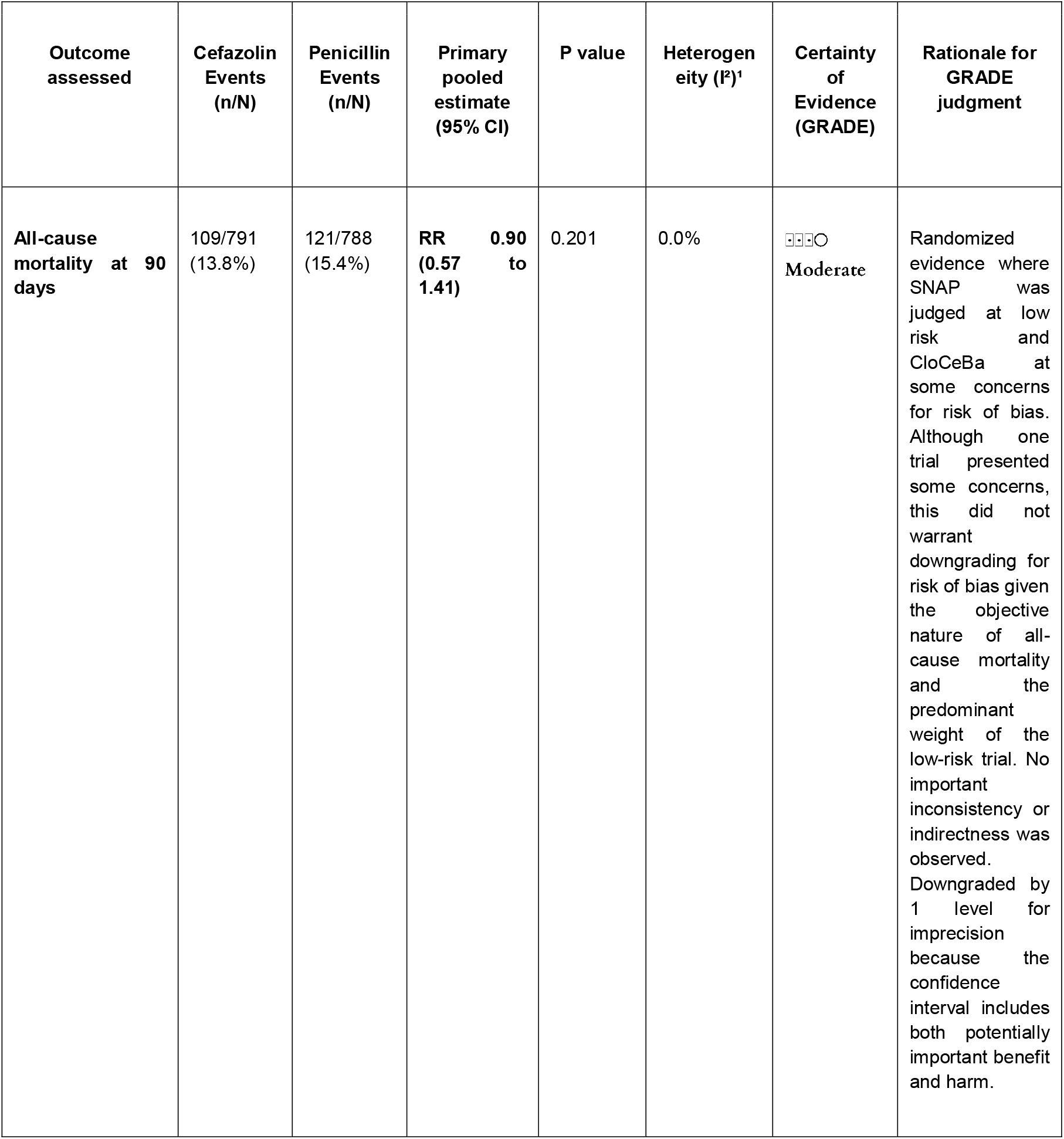
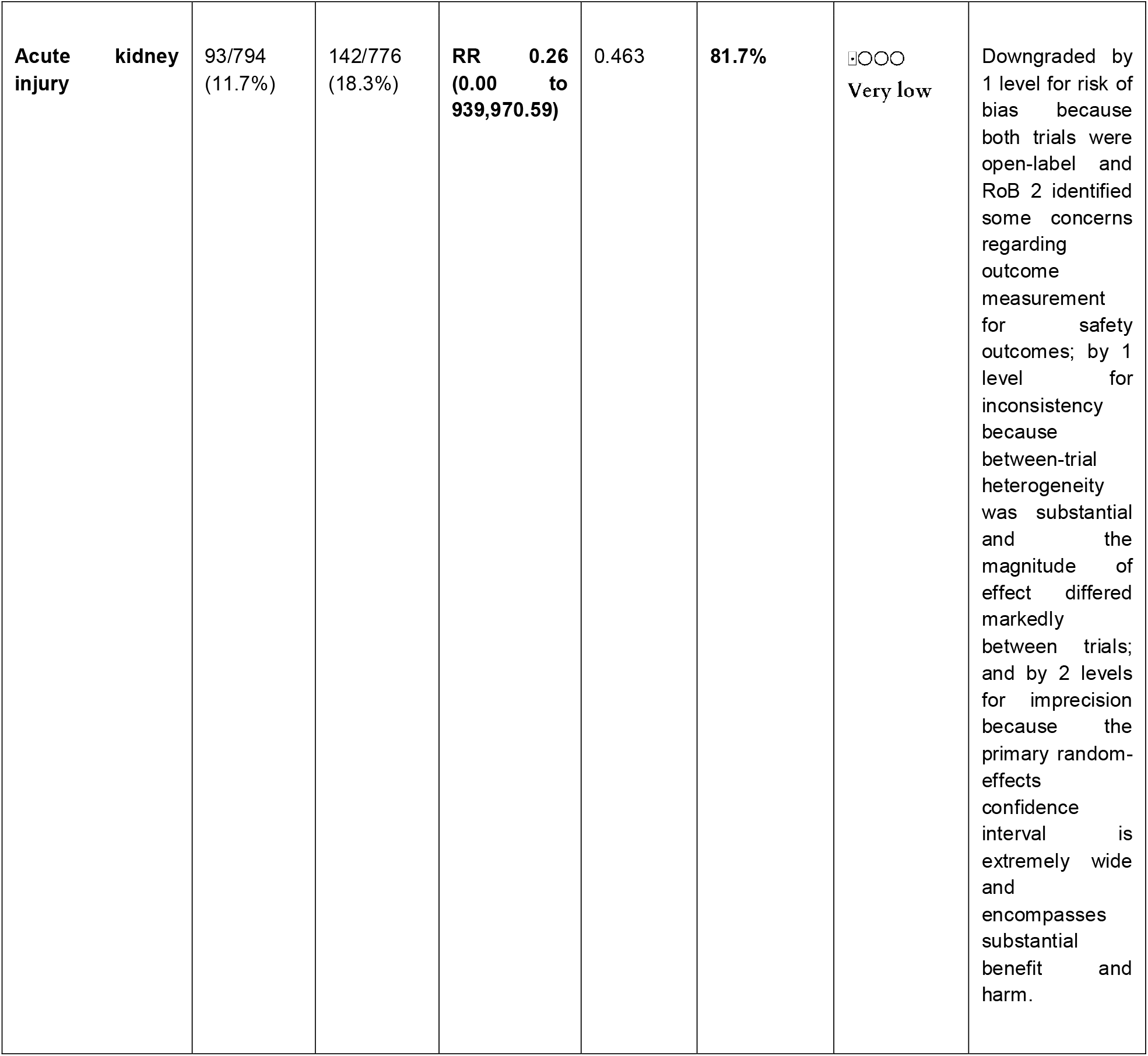
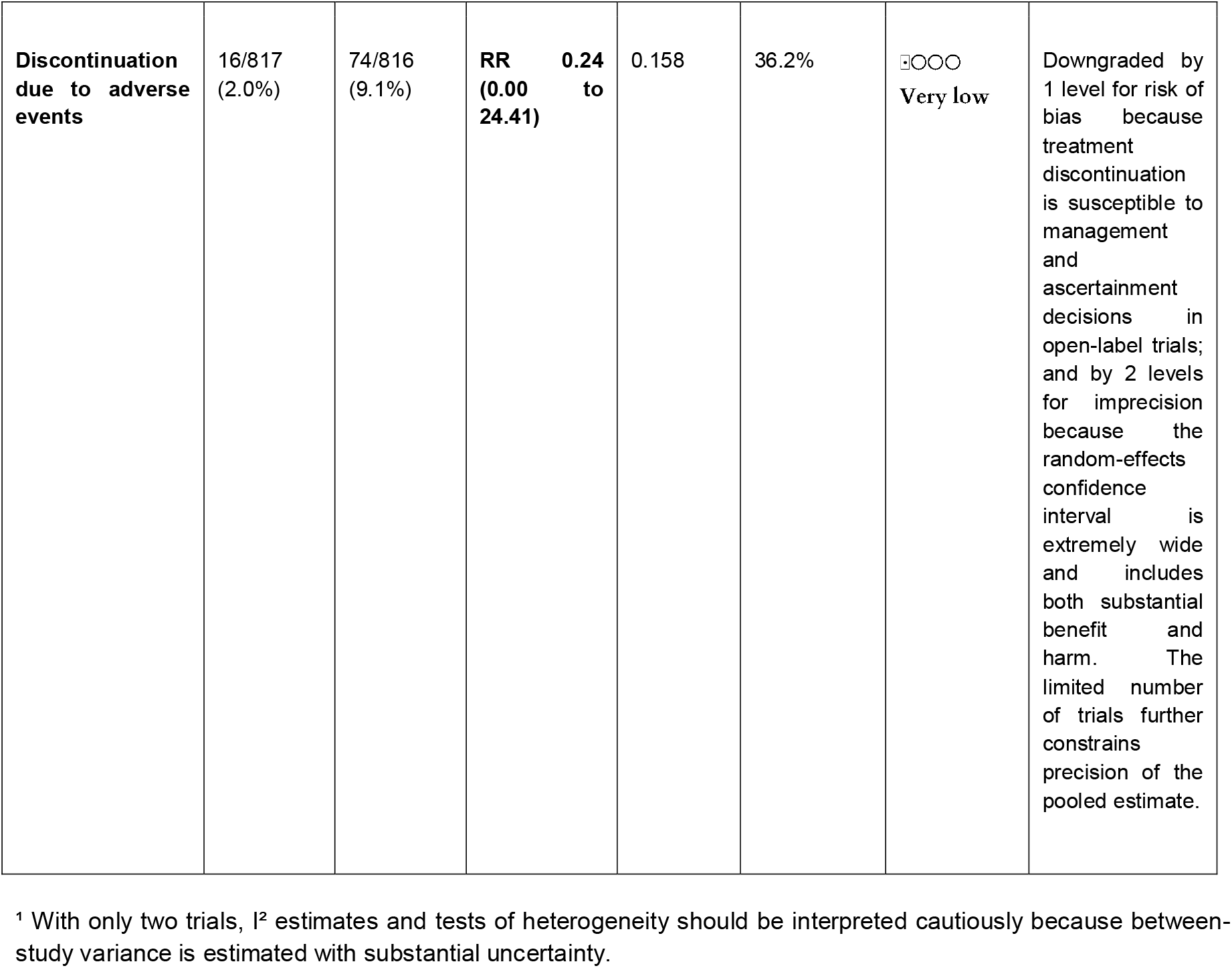
Summary of pooled effect estimates and certainty of evidence for mortality, safety and treatment discontinuation comparing cefazolin with antistaphylococcal penicillins in adults with methicillin-susceptible *Staphylococcus aureus* bacteremia.

**Table 4.** Sensitivity analyses assessing the robustness of comparative outcomes between cefazolin and antistaphylococcal penicillins in adults with methicillin-susceptible *Staphylococcus aureus* bacteremia across statistical models and assumptions about missing mortality data.

| Outcome | Analysis | Effect estimate (95% CI) | P value | I<sup>2</sup> |
| --- | --- | --- | --- | --- |
| **90-day all-cause mortality** | Primary: Random-effects, Paule-Mandel + HKSJ | **RR 0.90 (0.57 to 1.41)** | 0.201 | 0.0% |
|  | Sensitivity 1: All-missing-dead | RR 0.90 (0.63 to 1.30) | 0.177 | 0.0% |
|  | Sensitivity 2: All-missing-alive | RR 0.87 (0.50 to 1.54) | 0.204 | 0.0% |
|  | Sensitivity 3: Extreme worst-case for cefazolin | RR 1.12 (0.73 to 1.70) | 0.189 | 0.0% |
| **Acute kidney injury** | Primary: Random-effects, Paule-Mandel + HKSJ | **RR 0.26 (0.00 to 939,970.59)** | 0.463 | 81.7% |
|  | Sensitivity: Fixed-effect Mantel-Haenszel | RR 0.64 (0.50 to 0.82) | <0.001 | 81.7% |
|  | Sensitivity: Random-effects risk difference | RD -0.079 (-0.412 to 0.254) | 0.204 | 54.4% |
| **Discontinuation due to adverse events** | Primary: Random-effects, Paule-Mandel + HKSJ | **RR 0.24 (0.00 to 24.41)** | 0.158 | 36.2% |
|  | Sensitivity: Fixed-effect Mantel-Haenszel | RR 0.22 (0.13 to 0.37) | <0.001 | 36.2% |
|  | Sensitivity: Odds ratio | OR 0.22 (0.00 to 24.71) | 0.153 | 33.8% |

**Note:** Primary pooled estimates were obtained using random-effects models with Paule-Mandel estimation and Hartung-Knapp-Sidik-Jonkman inference. Fixed-effect Mantel-Haenszel estimates were conducted as sensitivity analyses to assess robustness to the choice of meta-analytic model. Risk-difference and odds-ratio analyses were conducted as additional effect-measure sensitivity analyses where appropriate. Given the small number of included trials, heterogeneity statistics should be interpreted cautiously.

**Figure 2.**
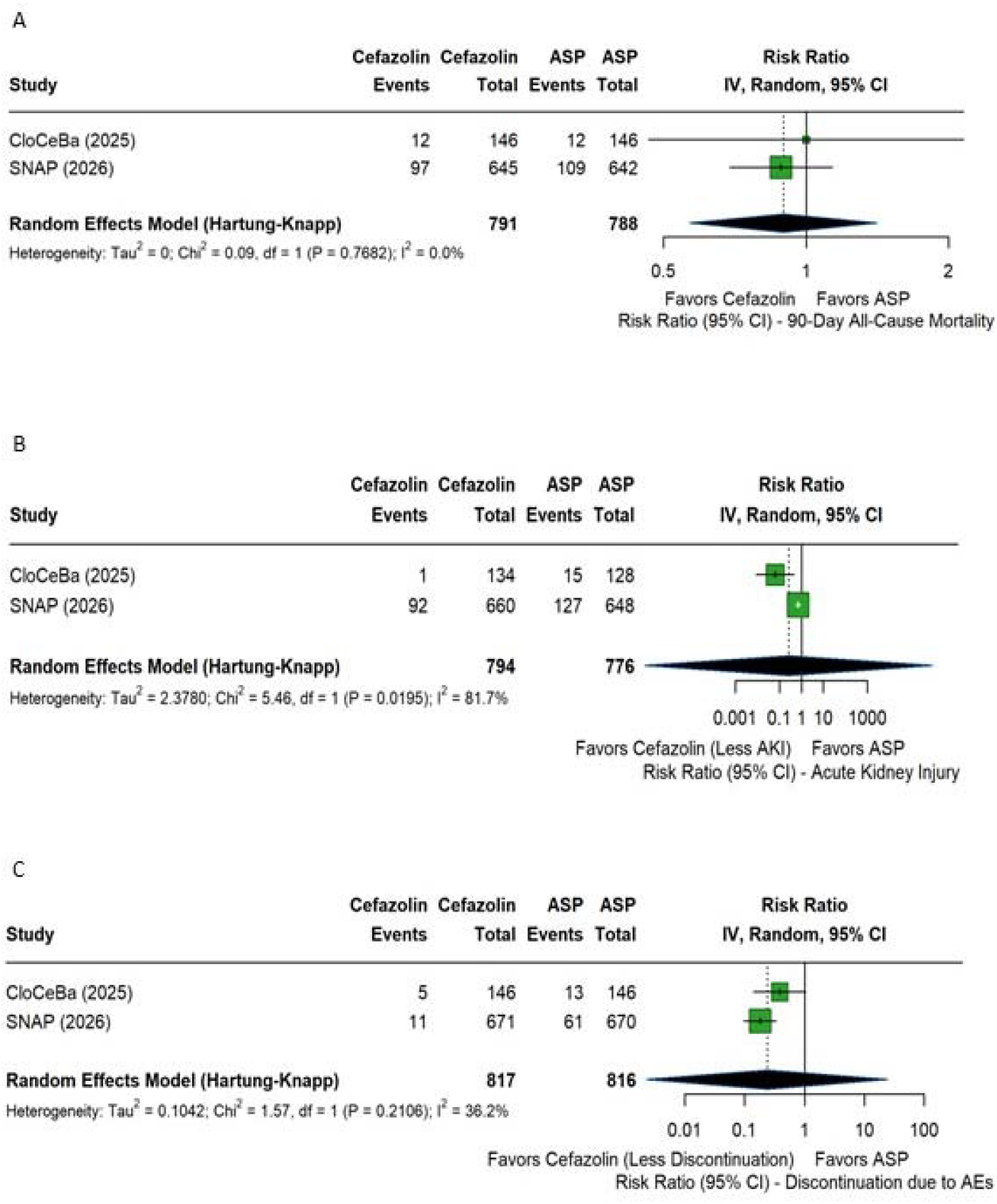
Forest plots of mortality, acute kidney injury and treatment discontinuation in randomized controlled trials comparing cefazolin with antistaphylococcal penicillins in adults with methicillin-susceptible *Staphylococcus aureus* bacteremia. (A) 90-day all-cause mortality; (B) acute kidney injury; and (C) treatment discontinuation due to adverse events.

#### 3.3.2. Primary Safety Outcome: Acute Kidney Injury (AKI)

Both trials favoured cefazolin, although effect magnitudes differed: AKI occurred in 13.9% versus 19.6% of participants in SNAP (RR 0.71) and 0.7% versus 11.7% in CloCeBa (RR 0.06) **(Figure 2B)**. The random-effects estimate was highly imprecise and did not reach statistical significance (RR 0.26, 95% CI 0.00-939,970.59; p = 0.463; I^2^ = 81.7%), whereas the fixed-effect estimate favoured cefazolin (RR 0.64, 95% CI 0.50-0.82; p < 0.001). Trial-level differences in AKI definitions and ascertainment, creatinine monitoring, baseline renal risk, eligibility criteria, and treatment regimens were examined descriptively **(Supplementary Table S3)**. These differences are plausible contributors to clinical and methodological heterogeneity, but their individual contributions cannot be determined from two trials. Absolute risk differences are presented as sensitivity analyses (**Table 4**).

#### 3.3.3. Treatment Discontinuation Due to Adverse Events

Treatment discontinuation due to adverse events occurred less frequently with cefazolin in both trials. The primary random-effects HKSJ analysis yielded an RR of 0.24 (95% CI 0.00-24.41; p = 0.158; I^2^ = 36.2%) **(Figure 2C)**. Although the direction of effect favoured cefazolin, the pooled estimate was highly imprecise. The fixed-effect Mantel-Haenszel sensitivity analysis produced a more precise estimate (RR 0.22, 95% CI 0.13-0.37), supporting the direction of the observed effect but not eliminating uncertainty related to the choice of model.

### 3.4. Non-Pooled Exploratory Outcomes

Few microbiological events were observed in either trial, however, outcomes were assessed over different time windows. In SNAP, microbiological treatment failure between days 15 and 90 occurred in 18/587 (3.1%) participants assigned to cefazolin and 12/566 (2.1%) assigned to antistaphylococcal penicillins. In CloCeBa, bacteremia relapse by day 90 occurred in 1/146 participants assigned to cefazolin and 2/146 assigned to cloxacillin. Because these outcomes differed in definition and assessment period, they were synthesized narratively rather than pooled.

Microbiological outcomes reflected three distinct aspects of infection control: early persistent bacteremia, relapse, and late microbiological failure. In CloCeBa, early bacteriological success was defined as negative blood cultures by day 3, or by day 5 in patients with infective endocarditis; persistent positive cultures indicated early persistent bacteremia, observed in 6.8% of cefazolin-treated participants vs 7.5% of the Cloxacillin group. Relapse was defined as a new MSSA-positive blood culture with the same antimicrobial susceptibility profile after documented early bacteriological success, occurring in 1/146 cefazolin-treated participants and 2/146 cloxacillin-treated participants. In SNAP, late microbiological failure referred to the reappearance of infection after treatment, assessed between days 15 and 90. This outcome occurred in 18/587 (3.1%) cefazolin-treated participants and 12/566 (2.1%) participants receiving antistaphylococcal penicillins (adjusted OR, 1.41; 95% CrI, 0.71-2.78). Neither trial reported a directly comparable 30-day mortality estimate, group-level data or hospital length of stay, not allowing quantitative synthesis.

## 4. DISCUSSION

### 4.1. Principal Findings

This systematic review and meta-analysis of two prospective RCTs, comprising 1,656 randomized participants, with 1,579 contributing to the pooled 90-day mortality analysis found no statistically significant difference in 90-day mortality between cefazolin and ASPs. Both trials reported fewer AKI events and treatment discontinuations with cefazolin; however, the magnitude and precision of the pooled safety estimates were sensitive to the meta-analytic model, particularly for AKI, for which substantial between-trial heterogeneity was observed. The random-effects HKSJ analyses therefore provide a more conservative representation of the uncertainty surrounding these effects. Fixed-effect sensitivity analyses yielded more precise estimates favouring cefazolin, but should not be interpreted as eliminating the underlying uncertainty. Overall, the available randomized evidence supports cefazolin as a reasonable alternative to antistaphylococcal penicillins, while the certainty regarding the magnitude of its safety advantage remains limited.

### 4.2. The Inoculum Effect and Clinical Outcomes

Historically, concerns regarding the clinical use of cefazolin have been driven in part by the in vitro cefazolin inoculum effect (CzIE), a laboratory phenomenon in which high bacterial densities and specific β-lactamase phenotypes can result in increased cefazolin hydrolysis in vitro [11]. Observational cohorts subsequently reported associations between the CzIE and therapeutic failure or persistent bacteremia [12], contributing to longstanding clinical reluctance to use cefazolin and reinforcing preferential use of ASPs.

The clinical significance of the cefazolin inoculum effect (CzIE) remains uncertain. Although observational studies have reported associations between the CzIE and adverse outcomes [12], neither included randomized trial was designed to determine whether this phenotype modifies the comparative effectiveness of cefazolin and antistaphylococcal penicillins. The individual trials did not provide a clear signal of worse overall clinical outcomes with cefazolin; however, microbiological endpoints differed between trials and were not pooled. These findings therefore neither refute the biological plausibility of the CzIE nor establish its clinical irrelevance, particularly in patients with high-inoculum infections or selected deep-seated foci.

### 4.3. Pathophysiological Mechanisms of Nephrotoxicity and Drug Tolerability

The lower AKI and treatment-discontinuation rates observed with cefazolin are biologically plausible, given the association of ASPs with interstitial nephritis and potential tubular toxicity [5,9]. In the pooled trials, AKI occurred in 11.7% (93/794) of cefazolin and 18.3% (142/776) of ASP recipients, and discontinuation due to adverse events in 2.0% (16/817) versus 9.1% (74/816), although the random-effects estimates were imprecise. However, AKI effects varied substantially between trials. Differences in renal eligibility, creatinine monitoring and treatment regimens may have contributed, but causation cannot be established **(Supplementary Table S3)**.

Hepatotoxicity did not differ in SNAP (13.1% versus 13.9%) [3]. Observational data suggest larger contrasts, including lower odds of nephrotoxicity (Peto OR 0.225), hepatotoxicity (0.160) and discontinuation (0.192) with cefazolin in hospitalized patients [9], and AKI in 19% with nafcillin versus 2% with cefazolin in a single-center cohort [26] (adjusted OR for nephrotoxicity 5.4 in another [27]). These contrasts likely reflect confounding by indication, heterogeneous definitions and a focus on nafcillin, and should not be taken as the expected effect size. Agent-level incidence could not be derived from the randomized evidence, as SNAP pooled flucloxacillin and cloxacillin, CloCeBa used cloxacillin, and neither trial evaluated nafcillin or oxacillin [3,4].

### 4.4. Implications for Clinical Practice and International Guidelines

These results provide randomized evidence relevant to the historical preference for ASPs in MSSA bacteremia. Cefazolin showed similar survival and microbiological outcomes while being associated with fewer toxicity-related treatment discontinuations. Its longer elimination half-life and less frequent intermittent dosing may also facilitate outpatient parenteral therapy [28,29].

Cefazolin therefore represents a reasonable option for many adults with MSSA bacteremia. Treatment should nevertheless remain individualized according to infection site, disease severity, source control, allergy history, organ function, and other clinical circumstances. The present evidence supports a reassessment of the historical hierarchy between cefazolin and ASPs rather than an assumption that one agent is universally preferable in all clinical scenarios.

### 4.5. Strengths and Limitations

Strengths include restriction to randomized evidence, which avoids the confounding by indication of observational comparisons; independent duplicate data extraction and RoB 2 assessment; extraction based on modified intention-to-treat populations; deterministic sensitivity analyses for missing mortality data; and a conservative primary analysis (random-effects, Paule-Mandel estimation, HKSJ inference), with fixed-effect, risk-difference and odds-ratio analyses as sensitivity checks. Our contribution is therefore not the identification of an additional randomized trial, but the quantitative appraisal of the consistency, precision and certainty of the randomized evidence.

Several limitations apply. First, only two trials (k = 2) were available, one contributing most participants. This precludes subgroup analyses, assessment of publication bias and reliable estimation of between-study variance. The HKSJ analysis yielded extremely wide confidence intervals for AKI, reflecting substantial uncertainty in random-effects inference with only two studies; the more precise fixed-effect estimate assumes a common underlying effect and should not be interpreted as resolving the observed heterogeneity. Second, AKI effects differed substantially in magnitude between trials despite a consistent direction of effect. Differences in renal eligibility criteria, creatinine assessment and available outcome data, treatment regimens, and comparator exposure may have contributed to this heterogeneity, but their individual contributions could not be disentangled. Third, both trials were open-label, potentially allowing differences in clinical management, adverse-event attribution, and treatment discontinuation. Although mortality is objective, the influence of treatment knowledge on safety outcomes cannot be excluded.

Fourth, microbiological outcomes were defined differently across trials and were therefore synthesized narratively. Fifth, no non-inferiority margin was prespecified for the pooled analysis, so the mortality estimate should not be interpreted as evidence of equivalence or non-inferiority. Sixth, few patients had high-inoculum infections or complicated left-sided endocarditis, patients with CNS infection were excluded, and neither trial evaluated nafcillin or oxacillin. Seventh, neither trial established whether phenotypically confirmed CzIE modifies clinical response. Finally, the search was expanded beyond PROSPERO-registered databases to improve completeness; this deviation is reported transparently.

### 4.6. Conclusions

In adults with MSSA bacteremia, available randomized evidence identified no statistically significant difference in 90-day all-cause mortality between cefazolin and antistaphylococcal penicillins. Cefazolin showed a directionally lower incidence of acute kidney injury and adverse-event-related treatment discontinuations, although statistical inference and precision for safety outcomes were highly model-dependent due to substantial between-trial heterogeneity and the limited number of studies. Microbiological outcomes were low-frequency events across trials and, due to heterogeneous definitions and assessment timeframes, could not be quantitatively pooled; narrative synthesis, however, showed no clear difference between groups. Overall, randomized trial evidence supports cefazolin as a reasonable definitive treatment option for MSSA bacteremia when appropriate clinical monitoring and source control are ensured, though the small number of trials warrants cautious interpretation.

## Data Availability

All data produced are available online at https://doi.org/10.5281/zenodo.23269884

## Acknowledgements

Frederico Larré Pozzebon acknowledges support received through a FAPERGS/PROBIC undergraduate research scholarship at Universidade Católica de Pelotas. This scholarship did not fund the present study and had no role in its design, conduct, analysis, interpretation, or decision to submit.

Ethics approval was not required for this systematic review and meta-analysis because the study used data from previously published and publicly available studies and did not involve the collection or analysis of individual-level data from human participants by the authors.

## Generative AI use

Generative Artificial Intelligence tools were used during manuscript preparation to assist with language refinement, organization of written content and text revision. All methodological decisions, data extraction, statistical analyses, interpretation of results, and final manuscript content were verified by the authors, who retain full responsibility for the accuracy, integrity and final version of the manuscript.

## Data availability

The aggregate data extracted from the included trials, the analytical dataset, and the R scripts used for the meta-analyses and sensitivity analyses are publicly available in Zenodo (DOI: https://doi.org/10.5281/zenodo.23269884). The repository includes documentation of variable definitions, software versions, and instructions for reproducing the analyses. All data were derived from published trial reports and their supplementary materials.

## Biographical note

**Frederico Larré Pozzebon** is a medical student at the Catholic University of Pelotas (UCPel), Brazil, and a recipient of a PROBIC/FAPERGS undergraduate research scholarship. He has contributed to scientific work presented at regional, national, and international conferences, including award-winning oral presentations. His academic interests include internal medicine, gut microbiota, and evidence-based clinical practice.

**Gustavo Treptow Cabral** is a medical student at the Catholic University of Pelotas (UCPel), Brazil. His academic interests include clinical medicine, scientific research, and evidence-based practice.

**Giovani Michelin Pacheco** is a medical student at the Catholic University of Pelotas (UCPel), Brazil. His academic interests include clinical medicine and evidence-based practice.

**Bibiana Vieira Dias** is a medical student at the Catholic University of Pelotas (UCPel), Brazil. Her academic interests include emergency medicine, general surgery, and evidence-based practice.

**Rodrigo Serra Macedo** is a medical student at the Catholic University of Pelotas (UCPel), Brazil. His academic interests include clinical medicine, dermatology, and scientific research.

**Adriano Martimbianco de Assis** holds a bachelor’s degree in Physical Education, a specialization in Health Sciences and Sports Medicine, and a master’s and doctoral degree in Biochemistry from the Federal University of Rio Grande do Sul (UFRGS), Brazil. He completed postdoctoral training at the Federal University of Health Sciences of Porto Alegre and UFRGS, and a research visit at the University of Lausanne, Switzerland. He is currently an Assistant Professor at the Catholic University of Pelotas (UCPel), where he coordinated the Graduate Program in Health and Behavior from 2021 to 2026. His academic expertise includes biochemistry, human physiology, cellular biology, neuroenergetics, and neuroprotection.

## Author Contributions

Frederico Larré Pozzebon: Conceptualization, Methodology, Investigation, Data curation, Formal analysis, Risk of bias assessment, Writing - original draft, Writing - review and editing, Project administration.

Gustavo Treptow Cabral: Investigation, Risk of bias assessment, Writing - original draft, Writing - review and editing.

Giovani Michelin Pacheco: Investigation, Data curation, Risk of bias assessment, Writing - original draft, Writing - review and editing.

Bibiana Vieira Dias: Writing - original draft.

Rodrigo Serra Macedo: Writing - review and editing.

Adriano Martimbianco de Assis: Conceptualization, Methodology, Writing - original draft, Writing - review and editing, Supervision.

All authors approved the final version of the manuscript and agreed to be accountable for all aspects of the work, ensuring that questions related to the accuracy or integrity of any part of the work are appropriately investigated and resolved.

## Image descriptions

**Figure 1. PRISMA 2020 flow diagram**. Flow diagram showing the identification, screening, eligibility assessment, and inclusion of records in the systematic review of randomized controlled trials comparing intravenous cefazolin with antistaphylococcal penicillins for methicillin-susceptible *Staphylococcus aureus* bacteremia in adults. The diagram presents the numbers of records identified, duplicates removed, records screened, reports assessed for eligibility, exclusions with reasons, and studies included in the qualitative and quantitative syntheses.

**Figure 2. Forest plots of efficacy and safety outcomes**. Forest plots comparing intravenous cefazolin with antistaphylococcal penicillins in adults with methicillin-susceptible *Staphylococcus aureus* bacteremia. The panels present pooled risk ratios and 95% confidence intervals for (A) 90-day all-cause mortality, (B) acute kidney injury, and (C) adverse event-related treatment discontinuation. Individual trial estimates and the pooled random-effects estimates are displayed, with the line of no effect at a risk ratio of 1. The plots illustrate the absence of a statistically significant mortality difference and the substantial uncertainty in the pooled safety estimates.

## References

1. Tong SYC, Davis JS, Eichenberger E, Holland TL, Fowler VG Jr. Staphylococcus aureus infections: epidemiology, pathophysiology, clinical manifestations, and management. Clin Microbiol Rev. 2015;28(3):603–661. doi:10.1128/CMR.00134-14.

2. Maraolo AE, Nobile M, Gentile I. Ceftriaxone for methicillin-susceptible Staphylococcus aureus bloodstream infections is associated with increased short-term mortality: a systematic review and meta-analysis. Ann Med. 2026;58(1):2667672. doi:10.1080/07853890.2026.2667672.

3. Lee TC, Barina LA, Walls G, Goodman AL, Yahav D, Cheng MP, et al.; Staphylococcus aureus Network Adaptive Platform (SNAP) Trial Group. Cefazolin for methicillin-susceptible Staphylococcus aureus bacteremia. N Engl J Med. 2026;394(23):2329–2339. doi:10.1056/NEJMoa2506905.

4. Burdet C, Saïdani N, Dupieux C, Lemaignen A, Canouï E, Surgers L, et al.; CloCeBa Study Group. Cloxacillin versus cefazolin for meticillin-susceptible Staphylococcus aureus bacteraemia (CloCeBa): a prospective, open-label, multicentre, non-inferiority, randomised clinical trial. Lancet. 2025;406(10517):2349–2359. doi:10.1016/S0140-6736(25)01624-1.

5. Perazella MA, Markowitz GS. Drug-induced acute interstitial nephritis. Nat Rev Nephrol. 2010;6(8):461–470. doi:10.1038/nrneph.2010.71.

6. Youngster I, Shenoy ES, Hooper DC, Nelson SB. Comparative evaluation of the tolerability of cefazolin and nafcillin for treatment of methicillin-susceptible Staphylococcus aureus infections in the outpatient setting. Clin Infect Dis. 2014;59(3):369–375. doi:10.1093/cid/ciu301.

7. Kwong JC, Urbancic K, Trubiano JA. Beyond penicillin: rapid desensitization for specific flucloxacillin hypersensitivity. Antimicrob Agents Chemother. 2018;62(12). doi:10.1128/AAC.01371-18.

8. Zand Irani A, Borchert G, Craven B, Gibbons H. Flucloxacillin and paracetamol induced pyroglutamic acidosis. BMJ Case Rep. 2021;14(1). doi:10.1136/bcr-2020-237536.

9. Eljaaly K, Alshehri S, Erstad BL. Systematic review and meta-analysis of the safety of antistaphylococcal penicillins compared to cefazolin. Antimicrob Agents Chemother. 2018;62(4). doi:10.1128/AAC.01816-17.

10. Schweizer ML, Furuno JP, Harris AD, Johnson JK, Shardell MD, Thom KA, et al. Comparative effectiveness of nafcillin or cefazolin versus vancomycin in methicillin-susceptible Staphylococcus aureus bacteremia. BMC Infect Dis. 2011;11:279. doi:10.1186/1471-2334-11-279.

11. Nannini EC, Stryjewski ME, Singh KV, Bourgogne A, Rude TH, Corey GR, et al. Inoculum effect with cefazolin among clinical isolates of methicillin-susceptible Staphylococcus aureus: frequency and possible cause of cefazolin treatment failure. Antimicrob Agents Chemother. 2009;53(8):3437–3441. doi:10.1128/AAC.00317-09.

12. Miller WR, Seas C, Carvajal LP, Diaz L, Echeverri AM, Ferro C, et al. The cefazolin inoculum effect is associated with increased mortality in methicillin-susceptible Staphylococcus aureus bacteremia. Open Forum Infect Dis. 2018;5(6). doi:10.1093/ofid/ofy123.

13. Livorsi DJ, Crispell E, Satola SW, Burd EM, Jerris R, Wang YF, et al. Prevalence of blaZ gene types and the inoculum effect with cefazolin among bloodstream isolates of methicillin-susceptible Staphylococcus aureus. Antimicrob Agents Chemother. 2012;56(8):4474–4477. doi:10.1128/AAC.00052-12.

14. Thomson KS. Extended-spectrum beta-lactamase, AmpC, and carbapenemase issues. J Clin Microbiol. 2010;48(4):1019–1025. doi:10.1128/JCM.00219-10.

15. Bourreau A, Le Mabecque V, Broquet A, Caillon J. Prevalence of a cefazolin inoculum effect associated with blaZ gene types, and clinical outcomes among methicillin-susceptible Staphylococcus aureus blood isolates of patients with infective endocarditis. Infect Dis Now. 2023;53(1):104626. doi:10.1016/j.idnow.2022.09.021.

16. Tsuji S, Gotoh K, Manabe T, Iio K, Fukushima S, Matsushita O, et al. Cefazolin inoculum effect in methicillin-susceptible Staphylococcus aureus clinical isolates. Diagn Microbiol Infect Dis. 2024;110(1):116399. doi:10.1016/j.diagmicrobio.2024.116399.

17. Goodman AL, Easom N, Bielopolski D, Sutherland RK, Marks M, Hensgens MPMH, et al. A review of the randomized clinical trial results from the Staphylococcus aureus network adaptive platform (SNAP) meticillin-susceptible (MSSA) and penicillin-susceptible (PSSA) domains and CloCeBa. J Antimicrob Chemother. 2026;81(8). doi:10.1093/jac/dkag215.

18. Page MJ, McKenzie JE, Bossuyt PM, Boutron I, Hoffmann TC, Mulrow CD, et al. The PRISMA 2020 statement: an updated guideline for reporting systematic reviews. BMJ. 2021;372. doi:10.1136/bmj.n71.

19. Higgins JPT, Thomas J, Chandler J, Cumpston M, Li T, Page MJ, et al., editors. Cochrane handbook for systematic reviews of interventions. Version 6.4. Cochrane; 2023. doi:10.1002/9781119536604.

20. Sterne JAC, Savović J, Page MJ, Elbers RG, Blencowe NS, Boutron I, et al. RoB 2: a revised tool for assessing risk of bias in randomised trials. BMJ. 2019;366. doi:10.1136/bmj.l4898.

21. IntHout J, Ioannidis JPA, Borm GF. The Hartung-Knapp-Sidik-Jonkman method for random effects meta-analysis is straightforward and considerably outperforms the standard DerSimonian-Laird method. BMC Med Res Methodol. 2014;14:25. doi:10.1186/1471-2288-14-25.

22. Balduzzi S, Rücker G, Schwarzer G. How to perform a meta-analysis with R: a practical tutorial. Evid Based Ment Health. 2019;22(4):153–160. doi:10.1136/ebmental-2019-300117.

23. Viechtbauer W. Conducting meta-analyses in R with the metafor package. J Stat Softw. 2010;36(3):1–48. doi:10.18637/jss.v036.i03.

24. Schwarzer G. meta: general package for meta-analysis [R package]. CRAN; 2026. doi:10.32614/CRAN.package.meta.

25. Guyatt GH, Oxman AD, Vist GE, Kunz R, Falck-Ytter Y, Alonso-Coello P, et al.; GRADE Working Group. GRADE: an emerging consensus on rating quality of evidence and strength of recommendations. BMJ. 2008;336(7650):924–926. doi:10.1136/bmj.39489.470347.AD.

26. Delgado V, Marsan NA, de Waha S, Bonaros N, Brida M, Burri H, et al. 2023 ESC Guidelines for the management of endocarditis. Eur Heart J. 2023;44(39):3948–4042. doi:10.1093/eurheartj/ehad193.

27. Norris AH, Shrestha NK, Allison GM, Keller SC, Bhavan KP, Zurawski CA, et al. 2018 Infectious Diseases Society of America clinical practice guideline for the management of outpatient parenteral antimicrobial therapy. Clin Infect Dis. 2019;68(1)–e35. doi:10.1093/cid/ciy745.

28. Chan L, Chan-Tompkins NH, Como J, Guarascio AJ. Retrospective analysis of adverse drug events between nafcillin versus cefazolin for treatment of methicillin-susceptible Staphylococcus aureus infections. Ann Pharmacother. 2020;54(7):662–668. doi:10.1177/1060028019897267.

29. Monogue ML, Ortwine JK, Wei W, Eljaaly K, Bhavan KP. Nafcillin versus cefazolin for the treatment of methicillin-susceptible Staphylococcus aureus bacteremia. J Infect Public Health. 2018;11(5):727–731. doi:10.1016/j.jiph.2018.02.004.

30. Buis DTP, Prins JM, Betica-Radic L, de Boer MGJ, Ekkelenkamp MB, van der Vaart TW, et al. Current clinical practice in antibiotic treatment of Staphylococcus aureus bacteraemia: results from a survey in five European countries. J Antimicrob Chemother. 2022;77(10):2827–2834. doi:10.1093/jac/dkac237.

31. Herrera-Hidalgo L, Muñoz P, Álvarez-Uría A, Alonso-Menchén D, Luque-Márquez R, Gutiérrez-Carretero E, et al. Contemporary use of cefazolin for MSSA infective endocarditis: analysis of a national prospective cohort. Int J Infect Dis. 2023;137:134–143. doi:10.1016/j.ijid.2023.10.019.

32. Nau R, Sörgel F, Eiffert H. Penetration of drugs through the blood-cerebrospinal fluid/blood-brain barrier for treatment of central nervous system infections. Clin Microbiol Rev. 2010;23(4):858–883. doi:10.1128/CMR.00007-10.

