## Supplementary table S1 for "Cefazolin versus Antistaphylococcal Penicillins for *Methicillin-Susceptible Staphylococcus aureus* Bacteremia: A Systematic Review and Meta-Analysis of Randomized Controlled Trials"

| **Database** | **Search Strategy / Syntax** | **Filters / Limits** | **Records Retrieved** |
| --- | --- | --- | --- |
| PubMed / MEDLINE | ("Cefazolin"[Mesh] OR cefazolin[tiab] OR cefazoline[tiab] OR cephazolin[tiab] OR cefamezin[tiab]) AND ("Staphylococcus aureus"[Mesh] OR "Bacteremia"[Mesh] OR "Staphylococcus aureus"[tiab] OR "S. aureus"[tiab] OR MSSA[tiab] OR SAB[tiab] OR "methicillin-susceptible Staphylococcus aureus"[tiab] OR "methicillin-sensitive Staphylococcus aureus"[tiab] OR bacteremia[tiab] OR bacteraemia[tiab] OR bloodstream infection*[tiab]) AND ("Oxacillin"[Mesh] OR "Nafcillin"[Mesh] OR "Cloxacillin"[Mesh] OR oxacillin[tiab] OR nafcillin[tiab] OR cloxacillin[tiab] OR flucloxacillin[tiab] OR antistaphylococcal penicillin*[tiab] OR beta-lactam*[tiab]) AND (randomized controlled trial[pt] OR controlled clinical trial[pt] OR randomi?ed[tiab] OR placebo[tiab] OR randomly[tiab]) NOT (animals[mh] NOT humans[mh]) | Humans | 22 |
| Embase | ('cefazolin'/exp OR cefazolin:ti,ab,kw OR cefazoline:ti,ab,kw OR cephazolin:ti,ab,kw) AND ('staphylococcus aureus'/exp OR 'bacteremia'/exp OR MSSA:ti,ab,kw OR SAB:ti,ab,kw OR bacteremia:ti,ab,kw OR bacteraemia:ti,ab,kw OR bloodstream infection*:ti,ab,kw) AND ('oxacillin'/exp OR 'nafcillin'/exp OR 'cloxacillin'/exp OR 'flucloxacillin'/exp OR oxacillin:ti,ab,kw OR nafcillin:ti,ab,kw OR cloxacillin:ti,ab,kw OR flucloxacillin:ti,ab,kw OR antistaphylococcal penicillin*:ti,ab,kw OR beta-lactam*:ti,ab,kw) AND ('randomized controlled trial'/exp OR 'controlled clinical trial'/exp OR 'randomization'/exp OR randomi?ed:ti,ab,kw) NOT ('animal'/exp NOT 'human'/exp) | Humans | 24 |
| Cochrane CENTRAL | ("Staphylococcus aureus" OR "S. aureus" OR MSSA OR SAB OR "bacteremia" OR "bloodstream infection*"):ti,ab,kw AND (cefazolin OR cephazolin OR cefamezin):ti,ab,kw AND (oxacillin OR nafcillin OR cloxacillin OR flucloxacillin OR "antistaphylococcal penicillin*"):ti,ab,kw | None (Trials registry) | 47 |
| Web of Science (Core Collection) | TS=(("Staphylococcus aureus" OR MSSA OR bacteremia OR "bloodstream infection*") AND (cefazolin OR cephazolin) AND (oxacillin OR cloxacillin OR flucloxacillin OR nafcillin OR "antistaphylococcal penicillin*") AND (randomized OR randomised OR "clinical trial" OR trial OR random*)) | None | 56 |
| Scopus | (cefazolin OR cefazoline OR cephazolin OR cefamezin) AND ("Staphylococcus aureus" OR "S. aureus" OR MSSA OR SAB OR "methicillin-susceptible Staphylococcus aureus" OR "methicillin-sensitive Staphylococcus aureus" OR bacteremia OR bacteraemia OR "bloodstream infection*") AND (oxacillin OR nafcillin OR cloxacillin OR flucloxacillin OR "antistaphylococcal penicillin*" OR "anti-staphylococcal penicillin*" OR "beta-lactam*") AND (random* OR randomised OR randomized OR randomization OR randomisation OR placebo OR "controlled trial" OR "clinical trial") | None | 423 |
| ClinicalTrials.gov | Condition: bacteremia due to staphylococcus aureus \| Intervention: cefazolin | — | 11 |
| Total | — | — | 583 |
