## Supplementary Table S2 for "Cefazolin versus Antistaphylococcal Penicillins for *Methicillin-Susceptible Staphylococcus aureus* Bacteremia: A Systematic Review and Meta-Analysis of Randomized Controlled Trials"

**Supplementary Table S2. Individual study weights in the random-effects meta-analyses**

| **Outcome** | **Study** | **Individual weight (%)** |
| --- | --- | --- |
| 90-day all-cause mortality (RR) | SNAP (2026) | 90.3 |
|  | CloCeBa (2025) | 9.7 |
| Acute kidney injury (RR) | SNAP (2026) | 58.9 |
|  | CloCeBa (2025) | 41.1 |
| Acute kidney injury (RD) | SNAP (2026) | 57.9 |
|  | CloCeBa (2025) | 42.1 |
| Treatment discontinuation due to adverse events (RR) | SNAP (2026) | 63.8 |
|  | CloCeBa (2025) | 36.2 |
| Late microbiological failure/recurrence (RR) | SNAP (2026) | 91.6 |
|  | CloCeBa (2025) | 8.4 |

Abbreviations: RR, risk ratio; RD, risk difference; SNAP, Staphylococcus aureus Network Adaptive Platform trial; CloCeBa, cefazolin versus cloxacillin trial.

Note: Study weights were obtained from the random-effects meta-analysis outputs using the inverse-variance method, with between-study variance estimated using the Paule-Mandel method and confidence intervals calculated using the Hartung-Knapp adjustment. Weights are reported separately for each outcome and effect measure; therefore, they should not be compared across different outcomes or effect measures as indicators of study quality.
