## Supplementary Table S3 for "Cefazolin versus Antistaphylococcal Penicillins for *Methicillin-Susceptible Staphylococcus aureus* Bacteremia: A Systematic Review and Meta-Analysis of Randomized Controlled Trials"

**Supplementary Table S3. Definitions, ascertainment, and treatment characteristics relevant to acute kidney injury in the SNAP and CloCeBa trials.**

| **Variable** | **SNAP trial** [3] | **CloCeBa trial** [4] |
| --- | --- | --- |
| Operational definition of AKI | AKI defined using serum creatinine changes according to the KDIGO classification. | AKI defined according to the KDIGO classification. |
| Laboratory criterion and creatinine threshold | Increase in serum creatinine of ≥0.3 mg/dL (≥26.5 μmol/L) or ≥50% from baseline. | KDIGO-based classification; the main article does not separately restate the numerical thresholds in its AKI results. |
| Baseline creatinine | Serum creatinine at study inclusion was used as the baseline for assessing subsequent changes. | Serum creatinine at inclusion was used as the reference for subsequent measurements. |
| Creatinine measurement schedule | Serum creatinine changes were assessed from inclusion to day 7 and to the end of study treatment, using available measurements. | Clinical and laboratory assessments were scheduled on days 1, 3, 5, and 7, at the end of study treatment, and at the end of antibiotic treatment. |
| AKI observation window | AKI assessed within 14 days; creatinine-based assessments included day 7 and end of study treatment. | AKI assessed using creatinine data collected during treatment; the principal reported comparison used day 7 measurements. |
| Ascertainment/adjudication method | Laboratory-based assessment using available serum creatinine measurements and the KDIGO classification. A separate independent AKI adjudication process is not specified in the main article. | Laboratory-based assessment using serum creatinine measurements and the KDIGO classification. A separate independent AKI adjudication process is not specified in the main article. |
| Population analyzed for AKI | Participants with available AKI outcome data: cefazolin, n = 660; (flu)cloxacillin, n = 648. | Participants with available day 7 creatinine data: cefazolin, n = 134; cloxacillin, n = 128. |
| AKI events, n/N (%) | Cefazolin: 92/660 (13.9%); (flu)cloxacillin: 127/648 (19.6%). | Cefazolin: 1/134 (0.7%); cloxacillin: 15/128 (11.7%). |
| Renal-function-related exclusion criteria | Patients receiving maintenance dialysis at enrolment were excluded. | Patients with severe renal impairment (eGFR below 30 mL/min/1.73 m²) were excluded. |

**Abbreviations**: AKI, acute kidney injury; ASP, antistaphylococcal penicillin; eGFR, estimated glomerular filtration rate; IV, intravenous; KDIGO, Kidney Disease: Improving Global Outcomes; MSSA, methicillin-susceptible Staphylococcus aureus.
