## Supplementary Table S4 for "Cefazolin versus Antistaphylococcal Penicillins for *Methicillin-Susceptible Staphylococcus aureus* Bacteremia: A Systematic Review and Meta-Analysis of Randomized Controlled Trials"

**Supplementary Table S4. Detailed RoB 2 signaling questions, supporting information, and judgments for the included randomized controlled trials.**

### **A. SNAP trial**

| **Domain / signaling questions** | **90-day all-cause mortality** | **Acute kidney injury (AKI) within 14 days** | **Antibiotic change due to adverse events** |
| --- | --- | --- | --- |
| D1. Randomization process | Low risk | Low risk | Low risk |
| 1.1 Random allocation sequence? | Y | Y | Y |
| 1.2 Allocation concealed until assignment? | Y | Y | Y |
| 1.3 Baseline differences suggest a problem? | N | N | N |
| Rationale | Simple 1:1 randomization (no stratification) through a web-based interactive system; the allocation was revealed only after eligibility was confirmed. No baseline imbalance suggesting a randomization problem (Table 1). | Same trial-level randomization process. | Same trial-level randomization process. |
| D2. Deviations from intended interventions | Low risk | Low risk | Low risk |
| 2.1–2.2 Participants and carers aware of assignment? | Y / Y | Y / Y | Y / Y |
| 2.3 Deviations arising because of trial context? | PN | PN | PN |
| 2.4–2.5 Deviations likely to affect outcome / balanced? | NA | NA | NA |
| 2.6 Appropriate analysis for effect of assignment? | Y | Y | Y |
| 2.7 Potential substantial impact from exclusions? | NA | NA | NA |
| Rationale | The trial was open-label, but awareness of allocation alone is not a deviation caused by the trial context. Changes of antibiotic for perceived inefficacy were uncommon and similar between groups (14/671, 2.1% vs 16/670, 2.4%), additional antibiotics after enrolment were discouraged, and the primary analysis followed randomized assignment regardless of receipt. | Same approach: no deviations arising from the trial context are described, and the analysis followed randomized assignment regardless of receipt. | Changes of antibiotic because of adverse events are the outcome itself, not deviations caused by the trial context; the susceptibility of this decision to awareness of allocation is addressed under D4. |
| D3. Missing outcome data | Low risk | Low risk | Low risk |
| 3.1 Data available for all or nearly all participants? | Y | Y | Y |
| 3.2 Evidence that missingness could not bias the result? | NA | NA | NA |
| 3.3 Could missingness depend on the true outcome value? | NA | NA | NA |
| 3.4 Is dependence on the true value likely? | NA | NA | NA |
| Rationale | The primary outcome was missing for 54/1,341 randomized participants (about 4%). Baseline characteristics were similar to those with complete data apart from younger age and injecting drug use, and the analyses assuming all missing participants alive or dead were congruent with the primary analysis (posterior probability of non-inferiority 99.3% and 99.6%). | AKI data were available for 660/671 (98.4%) and 648/670 (96.7%) participants. Even if all 11 missing cefazolin participants had AKI and none of the 22 missing comparator participants, AKI would be 103/671 (15.4%) vs 127/670 (19.0%), so the direction of the result is unchanged. | Denominators were 671 and 670 (11/671 vs 61/670), i.e. all randomized participants. |
| D4. Measurement of the outcome | Low risk | Some concerns | High risk |
| 4.1 Inappropriate measurement method? | N | N | N |
| 4.2 Could measurement differ between groups? | PN | PN | PN |
| 4.3 Outcome assessor aware of assignment? | Y | Y | Y |
| 4.4 Could knowledge influence measurement? | N | PY | Y |
| 4.5 Is influence likely? | NA | PN | PY |
| Rationale | All-cause mortality is objective and unlikely to be influenced by knowledge of allocation. | The creatinine-based definition was specified, but the frequency of creatinine monitoring was not collected. Differential testing therefore cannot be excluded, although the laboratory threshold itself is objective. | Whether to change the antibiotic because of an adverse event is a clinical decision made by unblinded clinicians. Knowledge of allocation, and the recognised tolerability concerns of antistaphylococcal penicillins, could influence attribution of events and the decision to stop or change treatment, and this influence cannot be separated from the observed difference (11/671 vs 61/670). |
| D5. Selection of the reported result | Low risk | Low risk | Low risk |
| 5.1 Result consistent with a prespecified analysis plan? | Y | PY | PY |
| 5.2 Multiple eligible outcome measurements? | PN | PN | PN |
| 5.3 Multiple eligible analyses? | PN | PN | PN |
| Rationale | Ninety-day mortality was the prespecified primary outcome, and the complete-case analysis and missing-data sensitivity analyses are described. | AKI was a specified secondary safety outcome, with a defined laboratory threshold. | Treatment change due to adverse events is a single binary outcome (Table 3), analysed with the same hierarchical Bayesian logistic model used for the other binary outcomes and reported once (aOR 0.21). No alternative definitions, time windows or analyses are described from which the result could have been selected. The outcome is not itemised in the main-text list of secondary outcomes, hence PY rather than Y for 5.1. |
| Overall risk of bias | Low risk | Some concerns | High risk |

### **B. CloCeBa trial**

| **Domain / signaling questions** | **90-day all-cause mortality** | **AKI at day 7** | **Premature discontinuation due to adverse events** |
| --- | --- | --- | --- |
| D1. Randomization process | Low risk | Low risk | Low risk |
| 1.1 Random allocation sequence? | Y | Y | Y |
| 1.2 Allocation concealed until assignment? | Y | Y | Y |
| 1.3 Baseline differences suggest a problem? | N | N | N |
| Rationale | Computer-generated blocks of 4, 6 or 8 prepared by an independent statistician, stratified by vascular-access-associated bacteremia and centre, with randomization performed in the electronic case report form to ensure allocation concealment. Baseline characteristics were balanced overall (mean Charlson score 4.4 vs 4.3); isolated differences (e.g. prosthetic joint 4% vs 12%) were judged compatible with chance. | Same trial-level randomization process. | Same trial-level randomization process. |
| D2. Deviations from intended interventions | Some concerns | Some concerns | Some concerns |
| 2.1–2.2 Participants and carers aware of assignment? | Y / Y | Y / Y | Y / Y |
| 2.3 Deviations arising because of trial context? | PN | PN | PN |
| 2.4–2.5 Deviations likely to affect outcome / balanced? | NA | NA | NA |
| 2.6 Appropriate analysis for effect of assignment? | PN | PN | PN |
| 2.7 Potential substantial impact from excluding randomized participants? | PN | PN | PN |
| Rationale | The trial was open-label. Of 315 randomized participants, 146 per arm were analysed: 12 of 158 (cefazolin) and 11 of 157 (cloxacillin) were excluded after randomization (7.6% vs 7.0%) for secondary exclusion (5 vs 6; meningitis, brain abscess or polymicrobial infection diagnosed after randomization), wrong inclusion (3 vs 3), investigator decision (3 vs 0) and consent withdrawal (1 vs 2). The exclusions are few, balanced and mostly eligibility-driven, so a substantial impact on the mortality estimate is unlikely (2.7 PN), but this is not a strict ITT analysis despite the authors' label, and the 3 unexplained investigator-decision exclusions occurred only in the cefazolin arm. Additional anti-staphylococcal antibiotics between day 2 and day 7 were more frequent with cloxacillin (21% vs 8%) but reflect clinical decisions during routine care rather than trial-context deviations (2.3 PN). | The same 23 post-randomization exclusions apply to the safety analysis. Even if all 12 excluded cefazolin participants had AKI and none of the 11 excluded cloxacillin participants, cefazolin would still have fewer events at day 7 (13/146 vs 15/139), so the impact is judged not substantial (2.7 PN). More frequent additional antibiotics with cloxacillin (21% vs 8%; aminoglycosides 6 vs 1) reflect clinical decisions during routine care rather than trial-context deviations (2.3 PN), and are noted as a limitation. | The same exclusions apply. They were few, balanced and unrelated to tolerability of the assigned drug, so a substantial impact on the discontinuation estimate is unlikely (2.7 PN), but the analysis is not a strict ITT. The higher use of additional antibiotics with cloxacillin is described but not classified as a deviation (2.3 PN). |
| D3. Missing outcome data | Some concerns | Some concerns | Low risk |
| 3.1 Data available for all or nearly all randomized participants? | PN | N | PN |
| 3.2 Evidence that missingness could not bias the result? | N | N | N |
| 3.3 Could missingness depend on the true outcome value? | PY | PY | PN |
| 3.4 Is dependence on the true value likely? | PN | PN | NA |
| Rationale | Survival was available for all 292 analysed participants (134/146 alive in each arm), but the 23 participants excluded after randomization (292/315 retained, 92.7%) contribute no data (3.1 PN), and no sensitivity analysis addressed them (3.2 N). Some exclusions (meningitis, brain abscess or polymicrobial infection) relate to illness severity, so missingness could depend on the true outcome (3.3 PY), but exclusions were few and balanced (12 vs 11), making an effect on the between-arm comparison unlikely (3.4 PN). | Day-7 creatinine was available for 134/146 cefazolin and 128/146 cloxacillin participants; relative to the 315 randomized, 53 lacked day-7 data (3.1 N). Missing values were not imputed and no sensitivity analysis was reported (3.2 N). Whether creatinine was repeated may depend on the clinical course (3.3 PY). However, even if all 12 missing cefazolin participants had AKI and none of the 18 missing cloxacillin participants, cefazolin would still have fewer events (13/146 vs 15/146), so bias is unlikely to explain the difference (3.4 PN). | The outcome was reported for all 292 analysed participants (5/146 vs 13/146); the 23 excluded participants contribute no data (3.1 PN) and no sensitivity analysis addressed them (3.2 N). The exclusion reasons are unrelated to tolerability of the assigned drug and were balanced between arms (12 vs 11), so missingness is unlikely to depend on the true outcome (3.3 PN). |
| D4. Measurement of the outcome | Low risk | Low risk | High risk |
| 4.1 Inappropriate measurement method? | N | N | N |
| 4.2 Could measurement differ between groups? | PN | PN | PN |
| 4.3 Outcome assessor aware of assignment? | Y | Y | Y |
| 4.4 Could knowledge influence measurement? | N | PN | Y |
| 4.5 Is influence likely? | NA | NA | PY |
| Rationale | Survival is an objective outcome. Open-label treatment alone does not imply biased measurement of death. | AKI was derived from serum creatinine (KDIGO classification) collected at protocol-scheduled visits (days 3, 5 and 7 and end of treatment) in both arms, an objective laboratory definition not plausibly influenced by knowledge of allocation, although investigators were unmasked. Lower availability of creatinine data with cloxacillin is addressed under D3. | Premature discontinuation due to an adverse event is itself a clinical decision made by unblinded investigators (the trial was open-label because of the different dosing schedules). Knowledge of allocation and the recognised tolerability concerns of cloxacillin could influence attribution and the decision to stop treatment, and this cannot be separated from the observed difference (5/146 vs 13/146). |
| D5. Selection of the reported result | Low risk | Some concerns | Low risk |
| 5.1 Result consistent with a prespecified analysis plan? | PY | N | PY |
| 5.2 Multiple eligible outcome measurements? | PN | PN | PN |
| 5.3 Multiple eligible analyses? | PN | PN | PN |
| Rationale | Survival at day 90 is a prespecified secondary outcome (the components of the composite primary outcome), measured at a single time point and reported once as a risk difference with its 95% CI. The protocol was published in advance and the trial is registered (NCT03248063). The primary analysis set presented is ITT rather than per protocol, following newer regulatory guidance; both are reported for the primary outcome. | The authors explicitly describe AKI as an additional, non-prespecified safety outcome (5.1 N). The standard KDIGO definition was used and both time points assessed (day 7 and end of study treatment) were reported in full, so there is no sign that a favourable definition, time point or analysis was selected (5.2 PN; 5.3 PN); selection cannot be fully excluded because the outcome was not prespecified. | Premature discontinuation due to an adverse event is listed among the prespecified safety outcomes (post hoc and non-prespecified safety outcomes are labelled separately), with a single definition and time frame, reported once, in a protocol published in advance. |
| Overall risk of bias | Some concerns | Some concerns | High risk |

**Abbreviations**: AKI, acute kidney injury; ITT, intention-to-treat; KDIGO, Kidney Disease: Improving Global Outcomes; NA, not applicable; NI, no information; PN, probably no; PY, probably yes; RoB 2, Cochrane Risk of Bias tool version 2; Y, yes.
